# Cardiometabolic biomarkers and systemic inflammation in US adolescents and young adults with latent tuberculosis infection: a population-based cohort study

**DOI:** 10.1101/2024.12.25.24319620

**Authors:** IM Magodoro, NAB Ntusi, J Jao, JZ Heather, BL Claggett, MJ Siedner, KA Wilkinson, RJ Wilkinson

## Abstract

**Background:** *Mycobacterium tuberculosis* (*M.tb*) infection in adults increases incident type 2 diabetes and atherosclerotic cardiovascular disease risk, but it is unknown if this cardiometabolic detriment occurs in the young. We sought to determine if young persons with latent tuberculosis infection (LTBI) have worse cardiometabolic health than their tuberculosis (TB) uninfected peers.

**Methods:** Peripubescent adolescents (12-15 years old) and older adolescents and young adults (16-30 years old) (older participants) were cross-sectionally surveyed. LTBI was assessed by tuberculin skin testing (induration ≥10mm). Fasting plasma glucose (FPG), HbA1c, c-peptide, NTproBNP, hs-Troponin T, c-reactive protein (CRP), ferritin, diabetes/prediabetes (FPG ≥5.6 mmol/L and/or HbA1c ≥5.7%) and homeostatic model of insulin resistance (IR) (HOMA2-IR) were measured as study outcomes. LTBI cases were propensity score matched 1:4 on sociodemographic and lifestyle indicators with TB uninfected controls to estimate adjusted median (adjMedian), mean differences (adjMD), and odds ratios (adjOR) of cardiometabolic indices.

**Results:** Seventy-five young persons with LTBI were matched with 300 TB uninfected peers of similar age [mean (SD): 18.3 (5.5) vs. 18.0 (5.5) years], race [Hispanic: 74.7% vs. 76.7%], and sedentary time [3.5 (1.5%) vs. 3.5 (1.6) hours/day]. LTBI was associated with higher inflammation [adjMedian (IQR) CRP: 0.22 (0.05, 0.34) vs. 0.11 (0.04, 0.35) mg/dL; p=0.027; ferritin: 55.0 (25.1, 90.3) vs. 41.1 (29.5, 136.2) ng/mL; p=0.047] among older participants, but not peripubescent adolescents [CRP: 0.08 (0.04, 0.36) vs. 0.05 (0.02, 0.17) mg/dL; p=0.42; ferritin: 23.0 (18.5, 33.5) vs. 32.7 (21.5, 48.2) ng/mL; p=0.011]. By contrast, there were no meaningful differences in FPG [adjMD (95%CI): −0.05 (−0.22, 0.12) mmol/L; p=0.57], HbA1c [0.0 (−0.17, 0.17) %; p=0.98] or diabetes/prediabetes prevalence [adjOR (95%CI): 0.9 (0.29, 2.29); p=0.85] by LTBI status. Insulin secretion and resistance, NTproBNP and hs-Troponin T were also similar.

**Conclusion:** Older adolescents and young adults with LTBI had greater markers of inflammation than those without LTBI while cardiometabolic profiles were similar across TB and/or age strata. Unlike in adults, *M.tb* infection in young persons does not appear associated with cardiometabolic derangement, although longterm consequences of chronic inflammation requires further study.

## Introduction

Active and latent (LTBI) infection with *Mycobacterium tuberculosis* (*M.tb*) in adults has been associated with both prevalent and new-onset disorders of cardiometabolism.^1, 2, 3, 4, 5^ Together with insights from mechanistic studies in humans and animals^6, 7, 8^, these observations have led to suggestions that tuberculosis (TB) might be a novel risk factor for type 2 diabetes mellitus (diabetes) and atherosclerotic cardiovascular disease (ASCVD). A diagnosis of TB in adults, for example, confers two-fold increased risk [relative risk (95%CI): 1.9 (1.4–2.7)] for incident cardiovascular (coronary, cerebrovascular, and peripheral arterial) morbidity and mortality according to a recent meta-analysis (2020)^1^; while prospective observational cohorts find up to five-fold increased risk for incident diabetes.^2, 4^ In human and murine tuberculosis, there is hypercoagulability^6^, elevated blood pressure^7^, endothelial and microvascular dysfunction, and cardiac hypertrophy and fibrosis^8^ and increased insulin resistance^3^, among others. These intermediate or preclinical forms are thought to be driven by an amplified and/or dysregulated inflammatory response to *M.tb* antigens.^9^

There are estimated to be 67 million adolescents (<15 years old) worldwide with LTBI (2014)^10^, and 1.8 million adolescents and young adults (10-24 years old) with incident active TB annually (2018) representing 17% of all new TB disease diagnoses globally.^11, 12^ If *M.tb* infection is cardiometabolically detrimental, this young population may be vulnerable to chronic disease and poor health outcomes in adulthood. *M.tb* exposure in early life will coincide with an accumulating burden of traditional risk factors like smoking, inadequate physical activity, excess calorie intake, and air pollution, among others.^13^ Although overt cardiometabolic disease is considered to affect only adults, it does have its beginnings in the first decade of life, and where its antecedents depend on the number, duration and intensity of risk factors.^14^

On the other hand, the inflammatory response to *M.tb* infection varies with age such that peripubescent children and adolescents have a balance of protective and tissue-damaging immunity compared to either younger (<c.5 years old) or older (>c.15 years old) people.^15^ Dubbed the “wonder years” of TB protective immunity, the risk of *M.tb* infection progressing to active TB disease in this age group is the lowest across the life span.^16, 17^ In the event of active TB, this period of life is classically characterized by paucibacillary, intra-thoracic disease with greater involvement of the mediastinal lymph nodes than the lung parenchyma.^17, 18, 19^ This is relatively clinically benign compared to the disseminated pathology which is more common in young children and the destructive pulmonary disease typical of adolescents and adults.^17, 18, 19^ However, the cardiometabolic correlates of this age-varying immunity to TB are unknown.

Therefore, understanding the nexus of *M.tb* infection, cardiometabolic health and age in young persons could be key to preventing TB-related diabetes and/or cardiovascular complications in adulthood as well as reducing their consequent burden for the future. Equally important, if this association is absent in young persons, it is imperative to identify and understand relevant protective factors. The objectives of our study, therefore, were to evaluate the hitherto under-examined cardiometabolic health of young persons (12-30 years old) with LTBI and how it varies with age, compared to a matched group of *M.tb* uninfected peers in the general population.

## Methods

We followed the guidelines for Strengthening the Reporting of Observational Studies in Epidemiology (STROBE)^20^ in the conduct and reporting of our analyses.

### Data Sources

Data for this study are derived from the National Health and Nutrition Examination Survey (NHANES), which are recurrent biennial cross-sectional examinations of the health status of children and adults in the US general population. Participants in the NHANES complete standardized questionnaires, physical examination, and laboratory testing of blood samples. Study protocols, including ethics and informed consent procedures, are described fully elsewhere.^21^ Because NHANES data are anonymized and publicly available, we did not seek institutional review board approval for the present study.

### Analytic Sample Selection

Our analysis was restricted to NHANES participants aged 12-30 years. Participants were excluded if they reported use of insulin and/or had missing data on tuberculin skin testing (TST) results, age, sex, race/ethnicity, household food security, physical activity, serum cotinine, and body mass index (**Figure 1**). These covariates were used for propensity score matching, as outlined below. We also excluded from the 1999-2000 NHANES cycle those noted to have Bacillus Calmette-Guérin (BCG) scars at the time of tuberculin skin testing. BCG vaccination status was not reported in the 2011-2012 NHANES cycle. We used the 1999-2000 NHANES cycle for the primary analysis and the 2011-2012 NHANES cycle for the secondary analysis.

**Figure 1.**
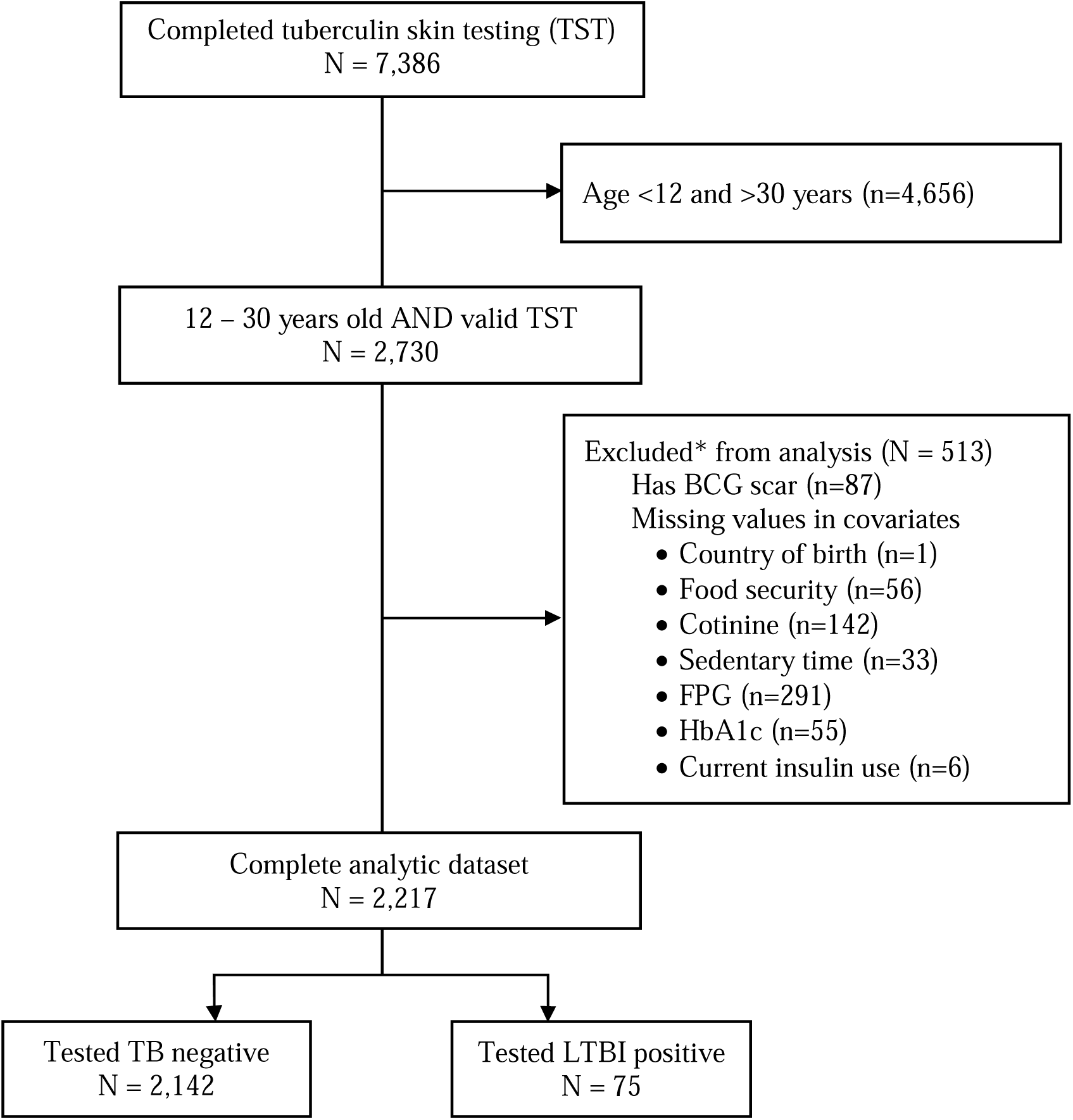
Flow chart of participant inclusion in final analytic sample using US NHANES 1999-2000.

### Study Measures

#### Latent tuberculosis infection

TST was undertaken using tuberculin-purified protein derivative (PPD) product, Tubersol® (Sanofi, Bridgewater, NJ). Skin induration was measured 48-72 hours after intradermal placement of PPD on the volar surface of the forearm with induration ≥10mm considered indicative of LTBI.^22^ Participants were also asked if they had ever had a TB (tine or tuberculin) skin test; were ever prescribed tuberculosis preventive therapy, if a positive TB skin test; ever had active TB; were ever prescribed antituberculosis drug treatment, if history of active TB; and ever lived in a household with an active TB contact. The presence of any one of these was taken to indicate likely prior TB exposure. However, chest radiographs were not collected nor were data on current tuberculosis symptoms. None of the participants were seropositive for HIV infection.

#### Cardiometabolic indices and inflammation markers

Fasting plasma glucose (FPG), fasting serum insulin and c-peptide concentrations were measured after an overnight fast of at least 9 hours, while glycated albumin (Alb1c) and hemoglobin (HbA1c), and lipid subfractions (HDL, LDL and triglycerides (TG)) were measured in nonfasting blood. NT-proBNP (limit of detection (LoD): ≥5pg/mL) (Roche Diagnostics, Indianapolis, IN), high-sensitivity troponin T (hs-cTntT) (LoD: ≥3 ng/L) (Roche Diagnostics, Indianapolis, IN), cystatin C (LoD, ≥0.23 mg/dL) (Dade Behring, Deerfield, IL), high-sensitivity C-reactive protein (hsCRP) (Dade Behring, Deerfield, IL) and ferritin (BioRad Laboratories, Hercules, CA) were measured in serum.^23^ The neutrophil-lymphocyte ratio (NLR) was calculated by dividing the absolute neutrophil count by the absolute lymphocyte count from a complete blood count (Beckman Coulter, Pasadena, CA). Participants also had their seated systolic and diastolic blood pressure (BP) measured four times using a mercury sphygmomanometer, of which we averaged the last three readings to obtain study BP values.

Homoeostasis model assessment (HOMA) 2 of insulin resistance (IR) (HOMA2-IR) was calculated using FPG and fasting c-peptide (or fasting insulin for the 2011-2012 NHANES) with the HOMA calculator (University of Oxford, Oxford, UK).^24^ Diabetes was defined as any of FPG ≥7.0 mmol/L or HbA1c ≥6.5%. Prediabetes was defined as any of FPG 5.6 to 6.9 mmol/L or HbA1c 5.7 to 6.4%.^25^ Because diabetes cases were too few for meaningful statistical inference, we merged diabetes and prediabetes to represent dysglycemia (diabetes/prediabetes). Data were not available to distinguish types 1 and 2 diabetes. However, we excluded all participants with self-reported current insulin thereby presumably excluding all type 1 diabetes cases.

#### Demographic, behavioral and biophysical measures

We categorized participants as peripubescent adolescents if 12-15 years old, and older adolescents and young adults (*i.e.,* older participants) if 16-30 years old. Sex was self-reported as either male or female. Participants described their race as any one of Hispanic, non-Hispanic white, non-Hispanic black, and non-Hispanic Asian/Other. We merged the latter three categories together as non-Hispanic. Country of birth was classified as either US or non-US. We assessed the socioeconomic status of participants by their household food security, which was reported as “food secure” or “food insecure”. Households were food insecure, and thus comparatively poor, if they reported ever relying on low-cost food, running out of food, skipping and/or reducing the size of meals at any point in the preceding 12 months. To assess physical (in)activity, we estimated sedentary time (hours/day) from participant reported time spent watching television, movies and/or using computer/games the previous day. We used serum cotinine, a nicotine biomarker, to assess tobacco smoke exposure from primary use as well as secondhand smoking.^26^ Weight and height (to calculate body mass index (BMI)) were also measured.

## Statistical Analysis

### Matching and covariate selection

To improve comparability and reduce bias, participants with LTBI were matched with TB uninfected peers using propensity scores (PS) matching. PS were estimated using multivariable logistic regression, with LTBI status as the outcome and the predictors as age, age group (12-15 years and 16-30 years), sex, race, country of birth, household food security (representing socioeconomic status), sedentary time, serum cotinine (representing smoking), and BMI. These covariates were selected as potential confounders of the LTBI/metabolism relationship or as risk factors for disordered metabolism. Matching was done 1:4 with nearest neighbor with no replacement within a specified caliper distance of 0.1. Between-group post-match balance was assessed by estimating the standardized mean difference (SMD) and the Kolmogorov-Smirnov (KS) statistic for each of the baseline covariates. An SMD <0.10 and KS <0.05 denote inconsequential residual bias.

Stratification by age group was informed by statistically significant tests evaluating the linear effects of LTBI status (*M.tb* uninfected vs. LTBI) on (i) hsCRP and (ii) ferritin incorporating interaction terms between TB status and age group (**Table S1**). Our *a priori* hypothesis was that *M.tb* related inflammation, and thus cardiometabolic risk, varies with age. FPG, Alb1c, HbA1c, HOMA2-IR, and dysglycemia prevalence, as indices of glucose metabolism, were our primary outcome measures. However, there were no significant *M.tb* status*age group interaction effects on any of these glucose metabolism indices (interaction term: p≥0.19) (**Table S1**) necessitating that only the main effects of LTBI are reported. For the secondary outcomes (detailed below) we nonetheless reported main effects of both LTBI and age group.

Differences and their 95% confidence intervals (95%CI) by LTBI status in the primary outcome measures were assessed through generalized linear models (GLM) using the identity link function for continuous outcomes and the logit link function for dichotomous outcomes. The GLM analyses were completed using (i) the unmatched sample without confounder adjustment (*unmatched*), and (ii) the PS matched sample without further confounder adjustment (*PS matched*). Differences in median NT-proBNP, hs-Troponin T, cystatin C, CRP, LDL, triglycerides and systolic BP by LTBI status and by age group were examined as secondary outcomes using quantile regression models in unmatched and PS matched samples. Their distributions were also visually compared using box-and-whisker plots. Lastly, descriptive statistics of study participants were presented stratified by LTBI status and according to variable scale as mean (SD), median (25^th^, 75^th^ percentile) or number (%).

### Secondary Analysis

We tested the consistency of our results in a different cohort collected a decade later, *i.e.*, 2011-2012 NHANES sample. We replicated the PS matching procedure using similarly defined covariates as in the primary analysis, and repeated the PS matched GLM analyses. Of note, BCG vaccine status was not reported in the 2011-2012 NHANES. Participants in the 2011-2012 NHANES cycle also completed an oral glucose tolerance test (OGTT) from which we obtained 2-hour post-load plasma glucose (PPG) in addition to FPG, fasting insulin, diabetes/prediabetes (FPG ≥5.6 mmol/L, or HbA1c ≥5.7% and/or PPG ≥7.8 mmol/L) and HOMA2-IR.

Analyses were conducted using R, version 3.6.3 (R Foundation for Statistical Computing, Vienna, Austria), and Stata version 17.0 (StataCorp, College Station, TX, USA), with Bonferroni-corrected p-values <0.007 considered statistically significant for the primary endpoints. Matching was performed using the MatchIt package^27^, and covariate balance was assessed using cobalt package^28^, both in R.

## Results

### Baseline characteristics of matched sample

The analytic sample consisted of 2,342 participants (**Figure 1**). PS matching achieved a balance of baseline covariates (**Table 1; Figure S1**) and yielded an effective sample size of 375, of which 75 had LTBI and 300 were *M.tb* uninfected controls. The mean (SD) age of participants in the matched sample was 18.3 (5.5) years for those with LTBI and 18.0 (5.5) years for the *M.tb* uninfected controls. The majority of participants were male (62.1%), of Hispanic race/ethnicity (76.3%) and were born outside the U.S (65.9%). Participants were of comparatively low socio-economic status given the common frequency of household food insecurity (33.1%). Although comparatively similar proportions of those with LTBI (72.6%) and their TB uninfected peers (66.0%; p=0.33) had a prior TB skin test, the former more frequently reported a positive test result (28.0% vs 4.3%; p<0.001) (**Table 2**). Overall, participants with LTBI (33.3%) were about five times as likely to have an indicator of likely prior TB exposure than TB uninfected controls (7.3%; p<0.001).

**Table 1.** Sociodemographic and behavioral characteristics of adolescent and young adult (12-30 years old) participants according to latent tuberculosis infection status before and after propensity score matching, US NHANES 1999-2000.

| Characteristic | Unmatched |  |  | PS matched |  |  |
| --- | --- | --- | --- | --- | --- | --- |
|  | *M.tb* uninfected | LTBI | SMD | *M.tb* uninfected | LTBI | SMD |
| Number | 2,142 | 75 |  | 300 | 75 |  |
| Mean age (years) | 17.8 (4.9) | 18.3 (5.5) | 0.087 | 18.0 (5.5) | 18.3 (5.5) | 0.050 |
| 12-15 years old | 961 (38.5%) | 32 (42.7%) | 0.085 | 136 (45.3%) | 32 (42.7%) |  |
| 16-30 years old | 1,534 (61.5) | 43 (57.3) |  | 164 (54.7%) | 43 (57.3%) | 0.054 |
| Female sex | 1,292 (51.8%) | 29 ( 38.7%) | 0.27 | 113 (37.7%) | 29 ( 38.7%) | 0.021 |
| Race/ethnicity |  |  |  |  |  |  |
| Hispanic | 1,100 (44.1%) | 56 (74.3%) | 0.66 | 230 (76.7%) | 56 (74.7%) | 0.070 |
| Non-Hispanic | 1,395 (55.9%) | 19 (25.3%) |  | 70 (23.3%) | 19 (25.3%) | 0.047 |
| Country of birth |  |  |  |  |  |  |
| USA | 442 (17.7%) | 25 ( 33.3) | 1.14 | 103 (34.3%) | 25 ( 33.3%) | 0.023 |
| Non-US | 2,053 (82.3%) | 50 (66.7%) |  | 197 (65.7%) | 50 (66.7%) |  |
| Household food security |  |  |  |  |  |  |
| Secure | 1,958 (78.5%) | 50 ( 66.7%) | 0.27 | 201 (67.0%) | 50 (66.7%) | 0.007 |
| Insecure | 537 (21.5%) | 25 (33.3%) |  | 99 (33.0%) | 25 (33.3%) |  |
| Sedentary time (hours/day) | 3.4 (1.6) | 3.5 (1.5) | 0.020 | 3.5 (1.6) | 3.5 (1.5) | 0.004 |
| Cotinine (ng/mL) | 0.16 (0.04, 1.98) | 0.05 (0.04, 1.13) | 0.27 | 0.08 (0.04, 1.06) | 0.05 (0.04, 1.59) | 0.075 |
| BMI (kg/m<sup>2</sup>) | 25.0 (6.3) | 24.6 (5.9) | 0.095 | 24.7 (5.6) | 24.6 (5.9) | 0.016 |

Values are mean (SD) or number (%) or proportion (%) or median (25<sup>th</sup>, 75<sup>th</sup> percentile).
PS = propensity score; SMD = standardized mean difference; BMI = body mass index.

**Table 2.** Tuberculosis-related history of unmatched and propensity score matched adolescent and young adult (12-30 years old) participants, US NHANES 1999-2000.

| Characteristic | Unmatched |  |  | PS matched |  |  |
| --- | --- | --- | --- | --- | --- | --- |
|  | *M.tb* uninfected | LTBI | p value | *M.tb* uninfected | LTBI | p value |
| Number | 2,142 | 75 |  | 300 | 75 |  |
| Tuberculin skin induration |  |  |  |  |  |  |
| Mean (SD) (mm) | 0.44 (1.25) | 13.9 (3.23) | <0.001 | 0.85 (1.89) | 13.9 (3.23) | <0.001 |
| Median (IQR) (mm) | 0.4 (0.0, 1.1) | 13.5 (12.0, 15.7) | <0.001 | 0.0 (0, 0) | 13.5 (12.0, 15.7) | <0.001 |
| Ever had TB skin test | 1,723 (71.5%) | 53 ( 72.6%) | 0.94 | 192 (66.0%) | 53 ( 72.6%) | 0.33 |
| Had negative TB skin test | 1,665 (66.7%) | 30 (40.0%) | <0.001 | 178 (59.3%) | 30 (40.0%) | <0.001 |
| Had positive TB skin test | 50 (2.0%) | 21 (28.0%) | <0.001 | 13 (4.3%) | 21 (28.0%) | <0.001 |
| Unknown/missing test result | 780 (31.3%) | 24 (32.0%) |  | 109 (36.3%) | 24 (32.0%) |  |
| Prescribed TBT | 29 (1.2%) | 17 (22.7%) | <0.001 | 9 ( 3.0%) | 17 (22.7%) | <0.001 |
| Ever had active TB | 5 (0.2%) | 2 ( 2.7%) | 0.004 | 2 (0.7%) | 2 (2.7%) | 0.041 |
| Prescribed ATT | 2 ( 0.1%) | 2 ( 2.7%) | <0.001 | 2 (0.7%) | 2 (2.7%) | 0.23 |
| Ever had household active TB contact | 44 (1.8%) | 8 (10.8%) | <0.001 | 10 (3.4%) | 8 (10.8%) | 0.001 |
| Any indicator of likely prior TB exposure<sup>a</sup> | 93 (3.7%) | 25 (33.3%) | <0.001 | 22 (7.3%) | 25 (33.3%) | <0.001 |

Values are mean (SD) or number (%) or median (25<sup>th</sup>, 75<sup>th</sup> percentile).
<sup>a</sup> Presence of any one of: (1) Ever had positive TB skin test; (2) Ever had active TB; (3) Ever prescribed ATT for active TB; and (4) Ever had household active TB contact.
PS = propensity score; LTBI = latent tuberculosis infection; TB = tuberculosis; TBT = tuberculosis preventive therapy; ATT = antituberculosis therapy.

### Distribution of cardiometabolic biomarkers

*M.tb* status*age group interaction effects post-match were statistically significant for hsCRP [β-coefficient (95%CI): 0.12 (0.01, 0.25); p=0.031] and ferritin [0.49 (0.03, 0.95); p=0.038], not NLR [-0.12 (−0.35, 0.10); p=0.29] (**Table S1**). As such, LTBI was associated with higher hsCRP [median (IQR): 0.22 (0.05, 0.34) vs. 0.11 (0.04, 0.35) mg/dL; p=0.027] and ferritin [55.0 (25.1, 90.3) vs. 41.1 (29.5, 136.2) ng/mL; p=0.047] compared to TB uninfected controls only among older participants. In peripubescent adolescents, those with LTBI had lower ferritin [23.0 (18.5, 33.5) ng/mL] than controls [32.7 (21.5, 48.2) ng/mL; p=0.011] but comparable hsCRP [0.08 (0.04, 0.36) vs. 0.05 (0.02, 0.17) mg/dL; p=0.42] (**Figure 2**). Excerpting higher LDL [101 (81, 124) vs. 86 (74, 108) mg/mL; p<0.001] and SBP [113 (104, 120) vs. 109 (104, 115) mmHg; p=0.011] in older participants than peripubescent adolescents, there were neither statistically nor clinically significant differences by *M.tb* status or age group in myocardial stress (NT-proBNP) or necrosis (hs-Troponin T), cardiovascular risk (cystatin C; systolic BP) or triglycerides (**Figure 2**).

**Figure 2.**
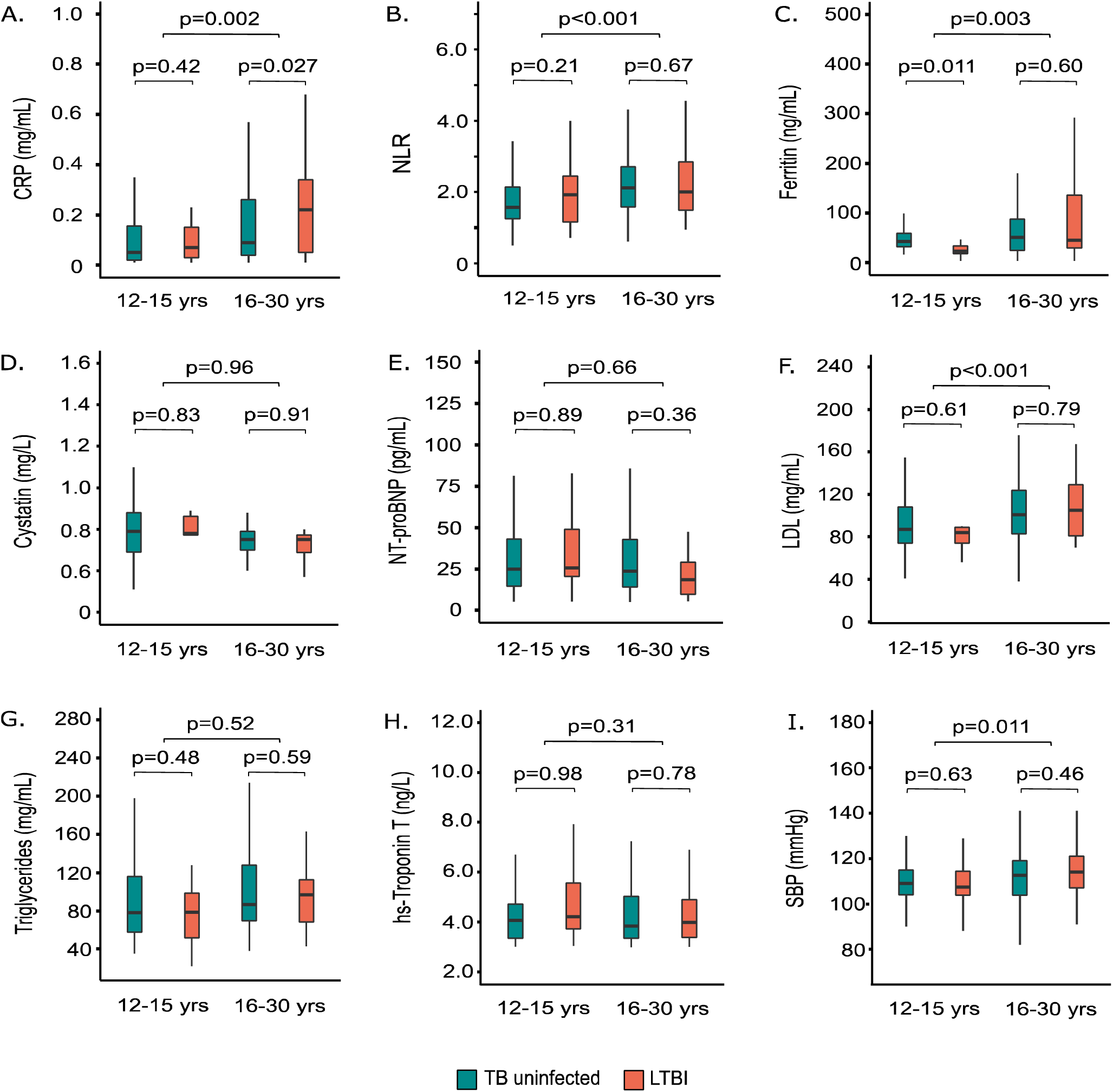
Cardiometabolic profiles of propensity score matched adolescent and young adult participants (12-30 years old) according to latent tuberculosis infection status and stratified by age, US NHANES 1999-2000. CRP = c-reactive protein; NLR = neutrophil-lymphocyte ratio; BNP = brain natriuretic peptide; HDL = high density lipoprotein cholesterol; SBP = systolic blood pressure; yrs = years old. p-values are for differences in median value using quantile regression models. Matched on age, sex, race, country of birth, household food, sedentary time, serum cotinine, and BMI.

### Glucose metabolism outcomes

The odds of having dysglycemia (diabetes/prediabetes) were similar with or without LTBI both before [odds ratio (OR): 1.29 (0.45, 2.95); p=0.59] and after [adjusted odds ratio (AOR): 0.9 (0.29, 2.29); p=0.85] PS matching (**Table 3**). Insulin secretion as measured by fasting c-peptide was similar between participants with LTBI and controls [adjusted mean difference (AMD): −0.05 (−0.18, 0.09) nmol/L; p=0.51], as was insulin resistance measured by HOMA2-IR [AMD: −0.11 (−0.38, 0.15); p=0.41]. We also did not find any statistically or clinically significant differences in immediate-[FPG AMD: −0.05 (−0.22, 0.12) mmol/L; p=0.57], medium-[Alb1c AMD: −1.5 (−0.56, 3.0]%; p=0.52] or long-range [HbA1c AMD: 0.0 (−0.17, 0.17)%; p=0.98] glycemia following PS matching. Comparisons in the unmatched sample found similar results (**Table 3**).

**Table 3.**
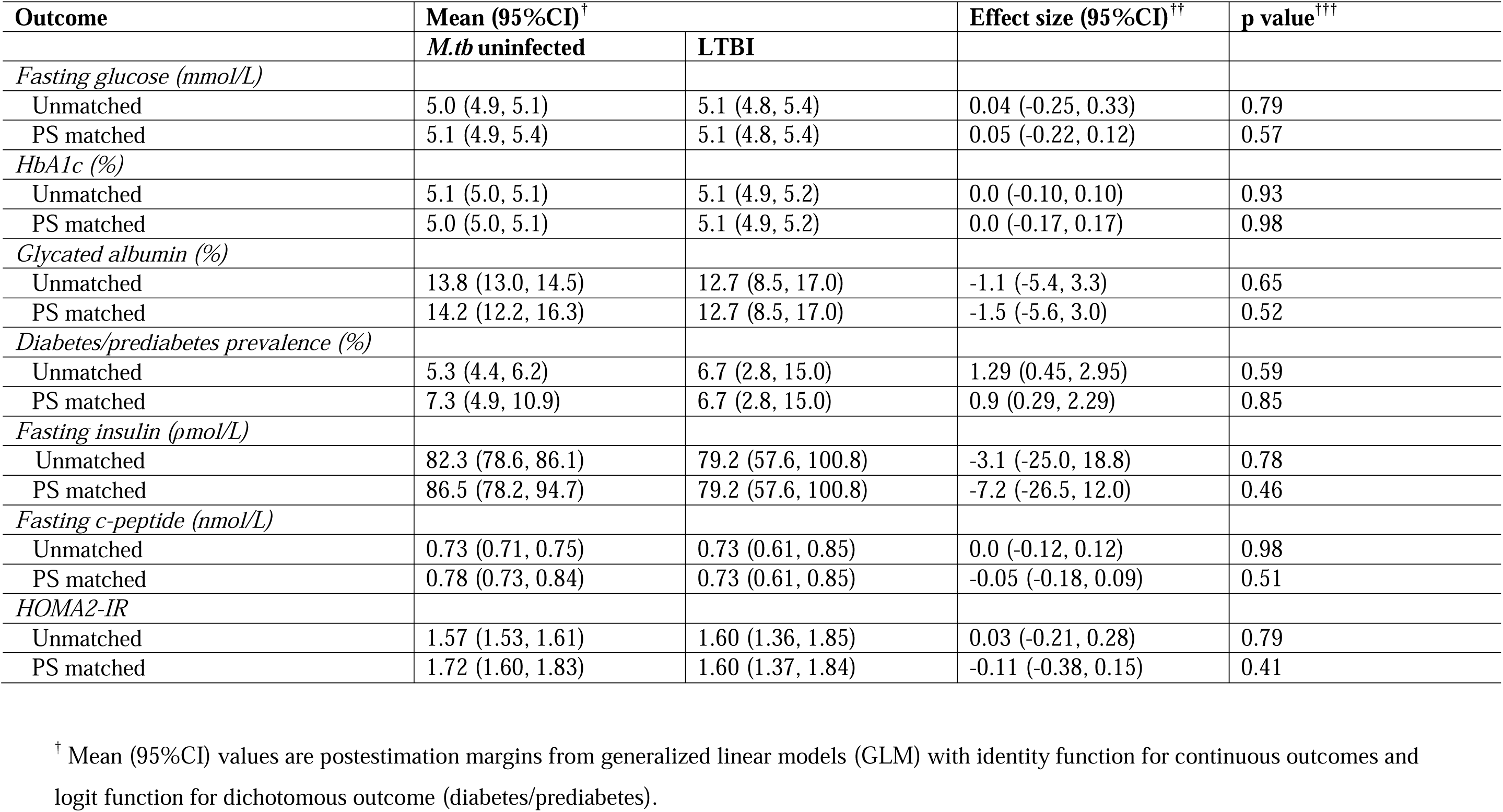
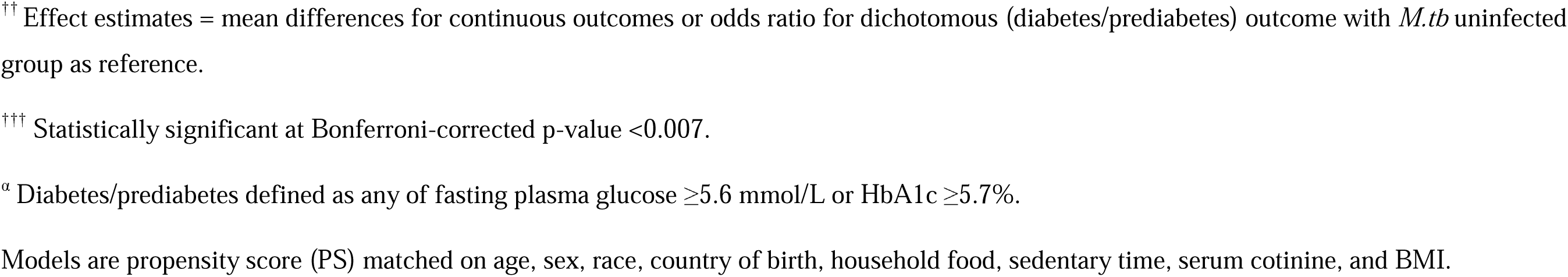
Associations between latent tuberculosis infection and glucose metabolism indices among PS matched adolescent and young adult participants (12-30 years old), unweighted US NHANES 1999-2000.

| Outcome | Mean (95%CI)<sup>†</sup> |  | Effect size (95%CI)<sup>††</sup> | p value<sup>†††</sup> |
| --- | --- | --- | --- | --- |
|  | *M.tb* uninfected | LTBI |  |  |
| *Fasting glucose (mmol/L)* |  |  |  |  |
| Unmatched | 5.0 (4.9, 5.1) | 5.1 (4.8, 5.4) | 0.04 (-0.25, 0.33) | 0.79 |
| PS matched | 5.1 (4.9, 5.4) | 5.1 (4.8, 5.4) | 0.05 (-0.22, 0.12) | 0.57 |
| *HbA1c (%)* |  |  |  |  |
| Unmatched | 5.1 (5.0, 5.1) | 5.1 (4.9, 5.2) | 0.0 (-0.10, 0.10) | 0.93 |
| PS matched | 5.0 (5.0, 5.1) | 5.1 (4.9, 5.2) | 0.0 (-0.17, 0.17) | 0.98 |
| *Glycated albumin (%)* |  |  |  |  |
| Unmatched | 13.8 (13.0, 14.5) | 12.7 (8.5, 17.0) | -1.1 (-5.4, 3.3) | 0.65 |
| PS matched | 14.2 (12.2, 16.3) | 12.7 (8.5, 17.0) | -1.5 (-5.6, 3.0) | 0.52 |
| *Diabetes/prediabetes prevalence (%)* |  |  |  |  |
| Unmatched | 5.3 (4.4, 6.2) | 6.7 (2.8, 15.0) | 1.29 (0.45, 2.95) | 0.59 |
| PS matched | 7.3 (4.9, 10.9) | 6.7 (2.8, 15.0) | 0.9 (0.29, 2.29) | 0.85 |
| *Fasting insulin (pmol/L)* |  |  |  |  |
| Unmatched | 82.3 (78.6, 86.1) | 79.2 (57.6, 100.8) | -3.1 (-25.0, 18.8) | 0.78 |
| PS matched | 86.5 (78.2, 94.7) | 79.2 (57.6, 100.8) | -7.2 (-26.5, 12.0) | 0.46 |
| *Fasting c-peptide (nmol/L)* |  |  |  |  |
| Unmatched | 0.73 (0.71, 0.75) | 0.73 (0.61, 0.85) | 0.0 (-0.12, 0.12) | 0.98 |
| PS matched | 0.78 (0.73, 0.84) | 0.73 (0.61, 0.85) | -0.05 (-0.18, 0.09) | 0.51 |
| *HOMA2-IR* |  |  |  |  |
| Unmatched | 1.57 (1.53, 1.61) | 1.60 (1.36, 1.85) | 0.03 (-0.21, 0.28) | 0.79 |
| PS matched | 1.72 (1.60, 1.83) | 1.60 (1.37, 1.84) | -0.11 (-0.38, 0.15) | 0.41 |

<sup>†</sup> Mean (95%CI) values are postestimation margins from generalized linear models (GLM) with identity function for continuous outcomes and logit function for dichotomous outcome (diabetes/prediabetes).
<sup>††</sup> Effect estimates = mean differences for continuous outcomes or odds ratio for dichotomous (diabetes/prediabetes) outcome with *M.tb* uninfected group as reference.
<sup>†††</sup> Statistically significant at Bonferroni-corrected p-value <0.007.
<sup>α</sup> Diabetes/prediabetes defined as any of fasting plasma glucose $\geq 5.6$ mmol/L or HbA1c $\geq 5.7\%$ .
Models are propensity score (PS) matched on age, sex, race, country of birth, household food, sedentary time, serum cotinine, and BMI.

### Secondary Analysis

When PS matching was repeated using the 2011-2012 NHANES cycle, we achieved a matched (secondary) sample of 345 total. Of these, 69 had LTBI and 276 were *M.tb* uninfected (**Table S2**). The secondary analytic sample (**Table S2**) was slightly older [mean age 21.6 years vs. 18.2 years] and had a higher proportion of non-US born participants [75.1% vs. 65.9%] than the primary sample (1999-2000 NHANES) (**Table 1**). Notwithstanding, the odds of diabetes/prediabetes in the secondary sample were similar [AOR: 1.19 (0.62, 2.19); p=0.59] for the two exposure groups (**Table 4**). Neither were there notable differences in any of FPG [AMD: −0.07 (−0.48, 0.34) mmol/L; p=0.75], PPG [AMD: −0.36 (−1.15, 0.44) mmol/L; p=0.38], HbA1c [AMD: −0.04 (−0.22, 0.12)%; p=0.57] or HOMA2-IR [AMD: −0.88 (−0.55, 0.38); p=0.73].

**Table 4.** Secondary analysis: Propensity score matched associations between latent tuberculosis infection and glucose metabolism indices among adolescent and young adult participants (12-30 years old), US NHANES 2011-2012.

| Outcome | Mean (95%CI)<sup>†</sup> |  | *Effect size (95%CI)<sup>††</sup> | P value<sup>†††</sup> |
| --- | --- | --- | --- | --- |
|  | *M.tb* uninfected | LTBI |  |  |
| Fasting glucose (mmol/L) | 5.4 (5.2, 5.5) | 5.3 (4.9, 5.7) | -0.07 (-0.48, 0.34) | 0.75 |
| Post-load glucose (mmol/L)<sup>α</sup> | 5.8 (5.4, 6.2) | 5.5 (4.7, 6.2) | -0.36 (-1.15, 0.44) | 0.38 |
| HbA1c (%) | 5.3 (5.2, 5.4) | 5.3 (5.1, 5.4) | -0.04 (-0.22, 0.12) | 0.57 |
| Fasting insulin (pmol/L) | 85.0 (72.1, 98.0) | 101.1 (74.9, 128.0) | 16.2 (-12.9, 45.4) | 0.28 |
| Diabetes/prediabetes prevalence (%)<sup>β</sup> | 20.3 (16.9, 25.4) | 23.2 (14.7, 34.5) | 1.19 (0.62, 2.19) | 0.59 |
| HOMA2-IR | 1.65 (1.44, 1.86) | 1.57 (1.14, 1.99) | -0.88 (-0.55, 0.38) | 0.73 |

<sup>†</sup> Mean (95%CI) values are postestimation margins from generalized linear models (GLM) with identity function for continuous outcomes and logit function for dichotomous outcome (diabetes/prediabetes) in PS matched sample without covariate adjustment.
<sup>††</sup> Effect estimates = mean differences for continuous outcomes or odds ratio for dichotomous (diabetes/prediabetes) outcome with *M.tb* uninfected group as reference.
<sup>β</sup> Diabetes/prediabetes defined as any of fasting plasma glucose $\geq 5.6$ mmol/L, or HbA1c $\geq 5.7\%$ and/or post-load glucose $\geq 7.8$ mmol/L.
Models are propensity score (PS) matched 1:4 on age, sex, race, country of birth, household food, sedentary time, serum cotinine, and BMI.<sup>α</sup> 2-hour plasma glucose following standard 75g glucose oral tolerance test.

## Discussion

We compared cross-sectional cardiometabolic biomarkers among young persons (12-30 years old) in the U.S. with LTBI versus their *M.tb* uninfected peers. We hypothesized that *M.tb* infection, by driving inflammation, is associated with worse profiles of glucose metabolism and cardiovascular status in an age-dependent manner. Unlike in older adolescents and young adults (16-30 years old) among whom LTBI was associated with worse inflammation, there was no evidence of increased inflammation with *M.tb* infection in peripubescent adolescents (12-15 years old). In fact, the acute phase reactant, ferritin, was reduced. We had insufficient data to assess diabetes/pre-diabetes, a relatively rare outcome in such young people. However, we did not find any clinically meaningful differences in the remainder of glucose metabolism readouts in relation to LTBI. These findings were replicated in the secondary analysis from a separate survey, underscoring their robustness. The biomarkers assessed in our study represent wide domains of cardiometabolic health such as myocardial stress and necrosis, cardiorenal status, lipid metabolism, blood pressure, short-, medium- and long-range glycemia, including glucose load handling, and insulin sensitivity. That latent *M.tb* infection in young persons may not be associated with cardiometabolic derangement requires confirmation in larger samples and especially in high TB burden disease settings. Whether this is also the case for active TB remains to be established.

*M.tb* infection in adults associates with transient impairment of glucose regulation and new-onset diabetes.^2, 4^ Transient hyperglycemia occurs during active TB and resolves with antitubercular drug treatment (ATT). It is debated whether this is just stress hyperglycaemia^29^, as occurs in other severe systemic infections, or it reflects true metabolic dysfunction, especially when it outlasts ATT. Hyperglycemia not in the diabetic range persisting up to 4 months posttuberculosis has been reported in patients with pulmonary TB (PTB).^30^ Notably, these patients were not diabetic at the time of TB diagnosis or before. Regarding overt diabetes developing in the posttuberculosis period, it is also not clear whether this is presaged by any dysglycemia, transient or otherwise, during active TB.

A study using a UK nationwide primary care adult cohort reported adjusted incidence rate ratios (IRRs) for new-onset diabetes of 5.7 following PTB and 4.7 following extrapulmonary TB (EPTB) compared to the general population.^4^ Average posttuberculosis follow-up was 4 years. Another US-wide study found higher diabetes incidence in adults with reactive TST and/or IGRA than in non-reactive peers [adjusted HR (95% CI): 1.2 (1.2, 1.3)].^2^ Patients with diabetes incident within a 2-year lag time after TST and IGRA testing were excluded, ensuring diabetes was not prevalent and undiagnosed at time of LTBI diagnosis. The median follow-up period was 3 years. We are not aware of comparable studies in children and adolescents. Salindri *et. al.,* (2019) examined the Taiwan national health insurance database (2002– 2013), which covers 99.9% of the national population, and found a diabetes incidence rate of 0.25 (95%CI: 0.15, 0.43) per 1,000 person-years (pys) in young persons aged 15-24 years old previously treated for TB.^31^ Their study did not include an *M.tb* uninfected control group. However, diabetes incidence in a similar age group (10-24 years) in the general population in Taiwan was separately estimated to be 0.12 (0.11, 0.12) per 1,000 pys using the same national health insurance database (2000– 2009).^32^

*M.tb* infection results in the activation of pro-inflammatory cytokines, inflammasome activity, free oxygen radicals, advanced glycation end-products, leading to matrix destruction, cell death, fibrosis and ultimately disordered tissue remodeling. In the cardiovascular system, these drive insulin resistance, hypercoagulability, elevated blood pressure, and endothelial and microvascular dysfunction. These intermediate states are thought to mediate the link between TB and ASCVD in adults.^9, 33^ Cross-sectional studies in Saudi Arabia^34^, Egypt^35^, Uganda^36^, Peru^36^, and the US^37^ note increased odds of prevalent ASCVD with LTBI. A recent (2023) Canadian prospective cohort study (1985–2019) including nearly 50,000 adults followed for a median 19 years reported higher risk of incident ASCVD with LTBI compared to no TB [adjusted HR: 1.08 (0.99-1.18)].^38^ We did not, however, find any evidence that young persons with LTBI had worse profiles of myocardial stress (NT-proBNP) or necrosis (hs-cTnT), cardiorenal function (cystatin C) or blood pressure, for example, than their *M.tb* uninfected controls. Only systemic inflammation (hsCRP) was significantly higher among the older (16-30 years old) participants with LTBI. It is possible to hypothesize that chronically elevated hsCRP in those with LTBI may predispose them later in adulthood towards developing cardiovascular disease.

Although our findings require replication, we speculate about possible explanations. Puberty varies in age of onset, and the 12-15 years age band in our cohort may have spanned the peripubescent “wonder years” of protective immunity against TB.^15^ Because this age group effectively contains *M.tb* within granuloma, LTBI in these young persons is likely accompanied by optimal balance between pro- and anti-inflammatory responses.^39^ The reduced ferritin among peripubescent adolescents with LTBI in our study may reflect this. We thus postulate that this balanced immune state manifests as the lack of cardiometabolic derangement among our peripubescent adolescents. On the other hand, *M.tb* infection in older adolescents and adults is accompanied by amplified and/or dysregulated inflammatory responses to mycobacterial antigens with tissue destruction.^15, 16^ Indeed, we found worse inflammation with LTBI among our older participants (16-39 years old). This, however, was not accompanied by evidence of cardiometabolic detriment in this age group.

Future confirmatory work with larger samples and including both latent and (past) active TB is required. In TB endemic areas like sub-Saharan Africa (SSA) and Southeast Asia (SEA), pediatric and adolescent TB overlap with malnutrition and/or HIV. How, if, these TB comorbidities impact cardiometabolism also requires urgent attention. Coincidentally, these regions are emerging epicenters of premature onset adult diabetes and cardiovascular diseases.^40^ A wider array of immune biomarkers will help in elucidating potential protective mechanisms.^41^ Imaging modalities and physiological assessments like cardiac magnetic resonance (CMR) and pulse tonometry, for example, are sensitive measures of preclinical damage. Approaches combining these with immune-omics would shed light into both phenotypes and mechanisms of TB-related cardiometabolism in adolescents.

### Strengths and limitations

Our study is among the first to explore potential cardiometabolic detriment of LTBI in young persons. Participants of Hispanic race/ethnicity and non-US birth made up the majority of our sample, reflecting TB epidemiology in the US and having implications for the generalizability of our findings. Our study also has a number of limitations. Firstly, our sample size of persons with LTBI was small for a comparatively rare outcome like (type 2) diabetes in a young population like our study participants. As such we obtained imprecise confidence intervals [adjOR (95%CI): 0.94 (0.29, 2.29)] for prevalent dysglycemia (prediabetes and diabetes) with LTBI versus *M.tb* uninfected controls. This was similar [adjOR: 1.10 (0.62, 2.19)] in the secondary analysis using 2011-2012 cohort (versus the 1999/2000 primary analysis cohort). Because overt (type 2) diabetes is rare in this age cohort, we hypothesized that any *M.tb*-related deleterious cardiometabolic effects might instead manifest as intermediate substrates like worse FPG, HbA1c and Alb1c, and/or as suboptimal insulin sensitivity and glucose load handling, among others. Whereas FPG is immediate, Alb1c (2-3 weeks)^42^ and HbA1c (2-3 months) are markers of medium- and long-range glycemic control. We did not find any clinically meaningful differences in these indices by LTBI status either. Insulin secretion (assessed by fasting plasma c-peptide) and resistance (assessed by HOMA2-IR) were also similar, as was glucose tolerance. Another limitation was the use of surrogate biomarkers to represent cardiometabolic health. Imaging and physiological assessments would have been more specific and sensitive markers of preclinical cardiovascular disease. Data on TB-related symptoms, chest radiographs and sputum examinations in conjunction with TST would have enabled better stratification into those who have eliminated TB infection, controlled TB infection or had subclinical TB infection. Excluding participants with BCG scars ameliorated potential misclassification biases from false positive TST. Finally, TB and diabetes have a bidirectional relationship, and as a cross-sectional study, our results could reflect the impact of dysglycemia on TB, and limits the causal inferences that we can draw around directionality.

### Conclusion

Young persons with LTBI had higher levels of inflammation but similar cardiometabolic profiles compared to their TB uninfected peers. Unlike in adults, *M.tb* infection in young persons may not be associated with cardiometabolic derangement although the prognostic relevance of chronically elevated hsCRP remains uncertain. However, larger confirmatory studies are required, especially in SSA and SEA, as are detailed immune mechanistic studies with deep cardiometabolic phenotyping. These will be key to better understanding of the relationship between *M.tb* infection and risk of future cardiometabolic syndromes.

## Contributors

IMM, NABN, JJ, BLC, MJS, HJZ, KAW, RJW.

## Data sharing

Data are publicly available at https://www.cdc.gov/nchs/nhanes/index.htm. Programming code is available upon request.

## Data Availability

Data are publicly available at https://www.cdc.gov/nchs/nhanes/index.htm

## Supplementary Material

**Figure S1.**
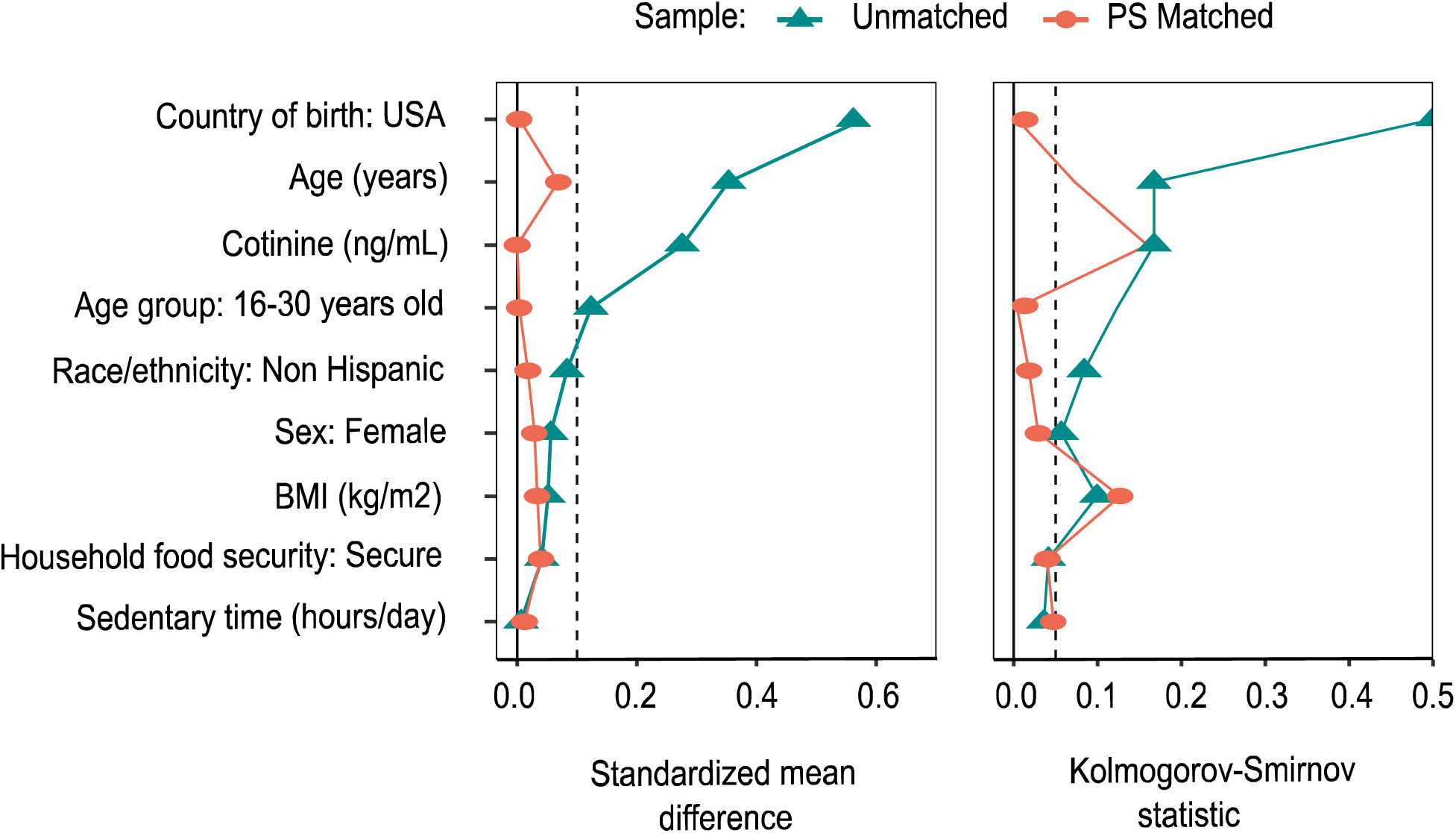
Covariate balance before and after propensity score matching.

**Table S1.** Main and interaction effects of age and latent tuberculosis infection on cardiometabolic indices among propensity score matched adolescents and young adults (12-30 years old), unweighted US NHANES 1999-2000.

| Biomarker | Effect estimates |  |  |  |  |  |
| --- | --- | --- | --- | --- | --- | --- |
|  | *Mtb* status<sup>†</sup> |  | Age group<sup>††</sup> |  | *Mtb* status*Age group<sup>†††</sup> |  |
| | $\beta$ (95%CI) | P value | $\beta$ (95%CI) | p value | $\beta$ (95%CI) | p value |
| Log hsCRP | 0.63 (0.08, 1.19) | 0.026 | 0.68 (0.35, 1.0) | <0.001 | 0.12 (0.01, 0.25) | 0.031 |
| Log NLR | 0.10 (-0.06, 0.27) | 0.23 | 0.27 (0.17, 0.37) | <0.001 | -0.12 (-0.35, 0.10) | 0.29 |
| Log Ferritin | -0.28 (-0.63, 0.07) | 0.12 | 0.37 (0.17, 0.58) | <0.001 | 0.49 (0.03, 0.95) | 0.038 |
| FPG (mmol/L) | -0.17 (-0.41, 0.07) | 0.16 | -0.13 (-0.27, 0.02) | 0.091 | 0.23 (-0.11, 0.57) | 0.19 |
| HbA1c (%) | -0.04 (-0.15, 0.07) | 0.44 | -0.04 (-0.10, 0.03) | 0.25 | 0.03 (-0.12, 0.17) | 0.71 |
| Alb1c (%) | -0.56 (-7.47, 6.28) | 0.87 | 1.54 (-2.58, 5.68) | 0.46 | -1.56 (-10.6, 7.52) | 0.73 |
| Fasting insulin (pmol/L) | -8.7 (-35.5, 18.0) | 0.52 | -19.4 (-35.8, -2.9) | 0.022 | -1.63 (-39.8, 36.5) | 0.93 |
| Fasting c-peptide (nmol/L) | -0.05 (-0.23, 0.12) | 0.55 | -0.03 (-0.14, 0.08) | 0.62 | -0.01 (-0.26, 0.25) | 0.97 |
| HOMA2-IR | -0.09 (-0.46, 0.28) | 0.64 | -0.01 (-0.59, 0.48) | 0.97 | -0.06 (-0.59, 0.48) | 0.84 |

<sup>†</sup> *Mtb* uninfected (reference group) versus LTBI.
<sup>††</sup> 12-15 years old (reference group) versus 16-30 years old.
<sup>†††</sup> *Mtb* status\*Age group = interaction term.
LTBI = latent tuberculosis infection; Log = natural logarithm; hsCRP = high sensitivity C-reactive protein; NLR = neutrophil lymphocyte ratio; FPG = fasting plasma glucose; Alb1c = glycated albumin; HbA1c = glycated hemoglobin; HOMA2-IR = homeostatic model assessment of insulin resistance.

**Table S2.** Characteristics of adolescent and young adult participants (12-30 years old) according to latent tuberculosis infection status before and after propensity score matching, US NHANES 2011-2012.

| Characteristic | Unmatched |  |  | PS matched |  |  |
| --- | --- | --- | --- | --- | --- | --- |
|  | *M.tb* uninfected | LTBI | SMD | *M.tb* uninfected | LTBI | SMD |
| Number | 1,590 | 69 |  | 276 | 69 |  |
| Age (years) | 19.6 (5.5) | 21.5 (5.4) | 0.35 | 21.9 (5.6) | 21.5 (5.4) | 0.068 |
| 12-15 years old | 473 (29.7%) | 12 (17.4%) | 0.29 | 47 (17.0%) | 12 (17.4%) |  |
| 16-30 years old | 1,117 (70.3%) | 57 (82.6%) |  | 229 (83.0%) | 57 (82.6%) | 0.010 |
| Female sex | 781 (49.1%) | 30 (43.5%) | 0.13 | 128 (46.4%) | 30 (43.5%) | 0.058 |
| Race/ethnicity |  |  |  |  |  |  |
| Hispanic | 420 (26.4%) | 24 (34.8%) | 0.18 | 101 (36.6%) | 24 (34.8%) | 0.048 |
| Non-Hispanic | 1,170 (73.6%) | 45 (65.2%) |  | 175 (63.4%) | 45 (65.2%) |  |
| Country of birth |  |  |  |  |  |  |
| USA | 1,311 (82.5%) | 17 (24.6%) | 1.42 | 69 (25.0%) | 17 (24.6%) | 0.008 |
| Non-US | 279 (17.5%) | 52 (75.4%) |  | 207 (75.0%) | 52 (75.4%) |  |
| Household food security |  |  |  |  |  |  |
| Secure | 1,172 (73.7%) | 48 (69.6%) | 0.092 | 203 (73.6%) | 48 (69.6%) | 0.088 |
| Insecure | 418 (26.3%) | 21 (30.4%) |  | 73 (26.4%) | 21 (30.4%) |  |
| Sedentary time (hours/day) | 3.0 (2.0, 4.0) | 3.1 (2.2, 4.3) | 0.007 | 3.0 (1.9, 4.1) | 3.1 (2.2, 4.3) | 0.012 |
| Cotinine (ng/dL) | 0.05 (0.01, 0.98) | 0.06 (0.02, 0.22) | 0.19 | 0.03 (0.01, 0.30) | 0.06 (0.02, 0.22) | <0.001 |
| BMI (kg/m<sup>2</sup>) | 25.6 (6.8) | 25.9 (6.6) | 0.051 | 25.7 (7.3) | 25.9 (6.6) | 0.032 |

Values are mean (SD) or number (%) or proportion (%) or median (25<sup>th</sup>, 75<sup>th</sup> percentile).
PS = propensity score; SMD = standardized mean difference; BMI = body mass index.
Matched on age, sex, race, country of birth, household food, LTPA, serum cotinine, and BMI.

